# Increased Risk for Type 2 Diabetes in Relation to Adiposity in Middle-Aged Black South African Men compared to Women

**DOI:** 10.1101/2021.10.19.21265228

**Authors:** Clement N. Kufe, Lisa K. Micklesfield, Maphoko Masemola, Tinashe Chikowore, Andre Pascal Kengne, Fredrik Karpe, Shane A. Norris, Nigel J. Crowther, Tommy Olsson, Julia H. Goedecke

## Abstract

**Aims:** Despite a higher prevalence of overweight/obesity in black South African women compared to men, the prevalence of type 2 diabetes does not differ. We explored if this could be due to sex differences in insulin sensitivity, clearance and/or beta-cell function, and also sex-specific associations with total and regional adiposity.

**Methods:** This cross-sectional study included 804 black South African men (n=388) and women (n=416). Dual-energy x-ray absorptiometry was used to measure total and regional adiposity. Insulin sensitivity (Matsuda index), secretion (C-peptide index) and clearance (C-peptide/insulin ratio) were estimated from an oral glucose tolerance test.

**Results:** After adjusting for sex differences in fat mass index, men were less insulin sensitive and had lower beta cell function than women (p<0.001), with the strength of the associations with measures of total and central adiposity being greater in men than women (p<0.001 for interactions). Further, the association between total adiposity and type 2 diabetes risk was also greater in men than women (relative risk ratio (95% confidence interval): 2.05 (1.42– 2.96), p<0.001 vs. 1.38 (1.03–1.85), p=0.031).

**Conclusion:** With increasing adiposity, particularly increased centralisation of body fat linked to decreased insulin sensitivity and beta cell function, black African men are at greater risk for type 2 diabetes than their female counterparts.

## Introduction

Type 2 diabetes (T2D) is a global health problem, with low-middle income countries particularly affected. It is projected that sub-Saharan Africa (SSA) will have the highest increase in T2D compared to the rest of the world, and in 2019 South Africa (SA) had the highest estimated number of people with diabetes (4.6 million) in the SSA region, and the highest age–adjusted comparative prevalence of diabetes (12.7%) in adults (1), which is higher than the global average (2). Within SSA and SA, the prevalence of T2D does not differ by sex, despite large sexual dimorphism in obesity rates (3). For example, in SA the prevalence of T2D in black SA men and women is similar (10.2% vs. 13.8%) (4), but the prevalence of overweight and obesity differs markedly (27.4% vs. 67.4%) (5).

The reason for this discrepancy in the association between overweight/obesity and diabetes risk in men and women is not clear. Our group have started to explore the underlying pathophysiology of T2D in Africans (6–10), and shown that black African women present with a phenotype of low insulin sensitivity and hyperinsulinemia due to higher insulin secretion and lower hepatic insulin clearance compared to white SA women (7) and black SA men (8). However, the majority of these studies have been undertaken in premenopausal women (6,7,10), with limited data in middle-aged men and women (8).

Notably, men typically have greater central fat mass (particularly visceral adipose tissue (VAT)) and less peripheral subcutaneous adipose tissue (SAT) than women, which is associated with a higher risk for T2D (8,11,12). However, the sex differences in the association between whole body and regional adiposity, and T2D risk, including insulin sensitivity, secretion and clearance, to our knowledge, has not been studied in African men and women.

Accordingly, the aims of this study were to compare insulin sensitivity, clearance and beta-cell function between middle-aged black South African men and women who differ in obesity prevalence, and to explore sex-specific associations with total and regional adiposity.

## Methods

This cross-sectional study includes the analysis of the follow-up data that was part of a longitudinal study designed to investigate the determinants of T2D risk in middle-aged black SA men and women. Data collection for the baseline study, as part of the AWI-Gen (Africa Wits-INDEPTH partnerships for Genomic Research) study (13), took place between 2011 and 2015 in black SA men (n=1027) and women (n=1008) residing in Soweto, South Africa (14). Follow-up data, analysed for this study, was collected between January 2017 and August 2018 on a sample of 502 men and 527 women randomly selected from the original sample. Participants living with HIV were excluded from this data analysis to avoid the confounding effects of the virus and antiretroviral therapy on the outcomes. Complete data was available for 804 participants (388 men and 416 women) and complete oral glucose tolerance (OGTT) data was available on 734 of these participants (Supplementary files: Figure 1).

**Figure 1:**
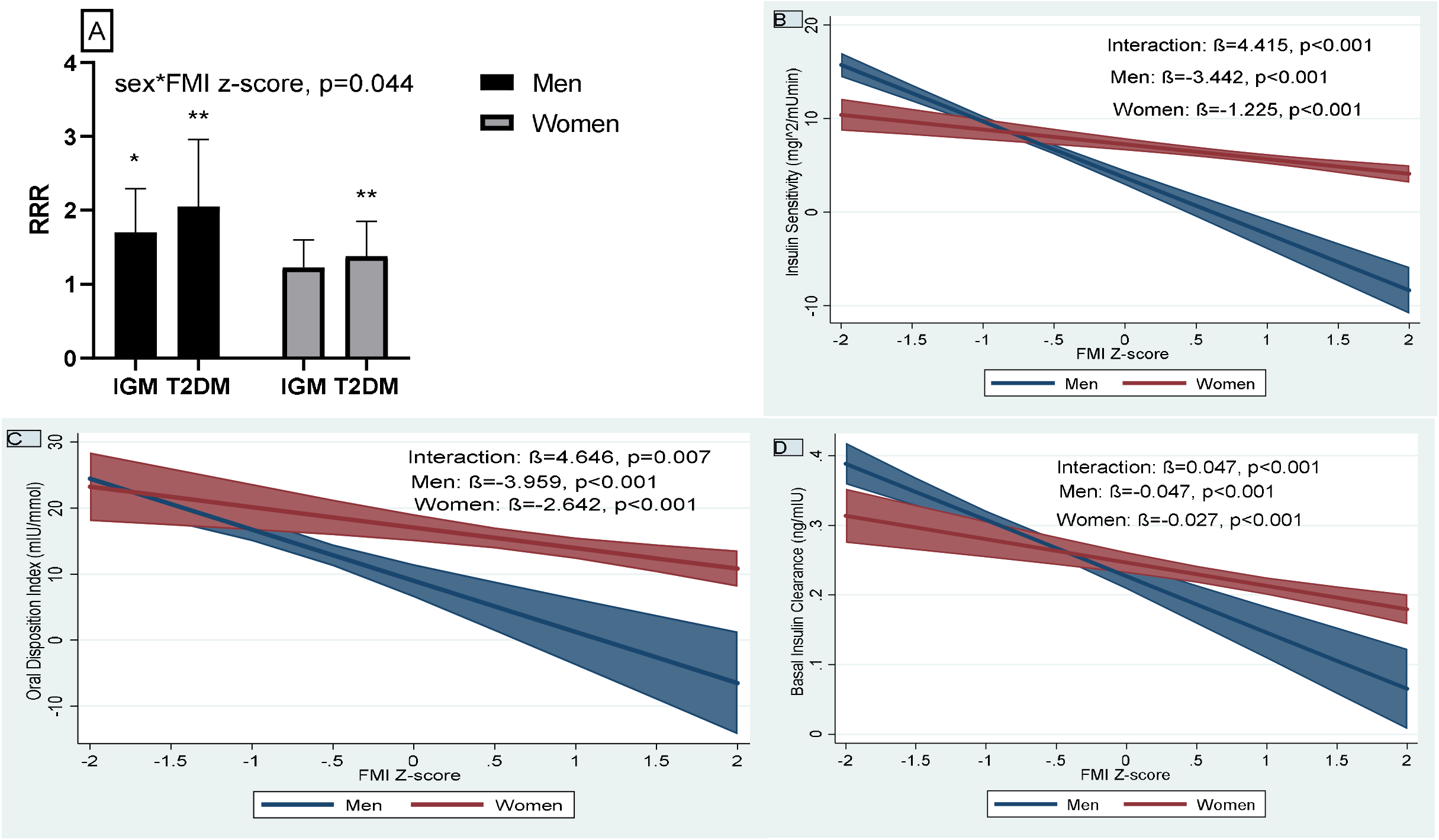
Bar Graph of the relative risk ratio (RRR) of impaired glucose metabolism (IGM) and type 2 diabetes mellitus compared to the normal glucose tolerant (NGT) in men and women, IGM: (RRR (95%CI): 1.70 (1.27–2.29), p<0.001 vs. 1.23 (0.95–1.60), p=0.115) and T2D: (2.05 (1.42–2.96), p<0.001 vs. 1.38 (1.03–1.85), p=0.031) for men and women, respectively (A); Sex-specific associations between FMI z-scores and insulin sensitivity (Matsuda index) (B), beta-cell function (oral disposition index) (C) and basal insulin clearance (D), modelled as predictive margins of sex with 95% CI

The study was conducted in accordance with the tenets of the Helsinki declaration and was approved by the Human Research Ethics Committee (HREC) of the University of the Witwatersrand (M160604 and M160975). Prior to inclusion in the study all procedures and possible risks were explained and all participants signed a consent form. Data collection took place at the South African Medical Research Council/University of the Witwatersrand Developmental Pathways for Health Research Unit at the Chris Hani Baragwanath Hospital in Soweto, Johannesburg, South Africa.

### Socio–demographic and medical questionnaire

Interviewer administered questionnaires were completed and captured onto REDCap (15). Data collected included age, marital status (married/unmarried), current employment (employed/not employed), highest educational level completed (no formal schooling/elementary school, secondary school level, tertiary education), alcohol intake and tobacco consumption (Yes/No), and self–reported diabetes and/or diabetes medication taken. Menopausal stage was classified according to last menstrual period (16).

### Anthropometry

Weight was measured to the nearest 0.1 kg using a TANITA digital scale (model: TBF-410, TANITA Corporation, US). Height was measured to the nearest 0.1cm using a wall–mounted stadiometer (Holtain, UK). Waist circumference (WC) and hip circumference (HC) were measured to the nearest 0.1cm with a non–stretchable tape. For the WC, the tape was placed horizontally between the iliac crest in the mid–axillary plane and the lowest rib margin. For the HC, the tape was placed around the level of the greatest protrusion of the buttocks. Waist-to-hip ratio (WHR) and BMI were calculated, and participants categorised according to the World Health Organisation (WHO) criteria (17).

### Body composition and body fat distribution measurements

Dual–energy X-ray absorptiometry (DXA) was used to measure whole body composition, including sub-total (total body minus head to account for any artefacts that may influence the DXA reading) fat mass (FM, kg and % body mass) and fat–free soft tissue mass (FFSTM), and regional FM including trunk, arm and leg FM (QDR 4500A, Hologic Inc., Bedford, USA, APEX software version 4.0.2). Fat mass index (FMI, sub-total fat mass kg/height^2^) and FFSTM index (FFSTM/height^2^) were calculated. Regional fat distribution was expressed relative to sub-total FM (%FM), with trunk fat (%FM) representing central fat distribution and arm and leg fat (%FM) representing upper- and lower-body peripheral fat distribution, respectively. Abdominal VAT and SAT areas were estimated from DXA (18).

### Blood sampling and analysis

Participants were instructed to not eat, smoke, drink alcohol or exercise for at least 8 hours prior to testing. A single baseline blood sample (10 ml) was drawn for the determination of glycated haemoglobin (HbA1c), plasma glucose, serum insulin, C–peptide and follicle stimulating hormone (FSH) concentrations. Participants then completed a standard 75g oral glucose tolerance test (OGTT) over 2-hours during which blood samples (5 ml) were drawn at 30 min intervals for the determination of glucose, insulin and C-peptide concentrations. Participants with known diabetes and/or those with fasting blood glucose ≥11.1 mmol/l (n=76) (ACCU-CHEK^®^, MedNet GmbH, Munster, Germany) did not complete the OGTT.

Plasma glucose concentrations were measured on the Randox RX Daytona Chemistry Analyser (Randox Laboratories Ltd., London, UK). HbA1c concentrations were measured using the D–10™ Haemoglobin Analyser (Bio–Rad Laboratories, Inc. USA). Serum insulin and C–peptide concentrations were measured on the Immulite® 1000 Immunoassay System (Siemens Chemiluminescent Healthcare GmbH, Henkestr, Germany). FSH was measured on serum using the ARCHITECT Chemiluminescent Microparticle Immunoassay assay (Abbott Laboratories, Abbott Ireland).

Based on the fasting plasma glucose (FPG) and 2–h OGTT glucose results, participants were classified according to the WHO criteria (19). Participants with impaired fasting glucose and impaired glucose tolerance were combined and described as having impaired glucose metabolism (IGM).

### Calculations from the OGTT

The homeostasis model assessment (HOMA-IR) was used to estimate fasting insulin resistance (20). The Matsuda Index (21), was used to estimate insulin sensitivity for participants with complete OGTT data (n=628), alongside the composite score (22) for participants who only had data for 0 and 120 minutes (n=106). These composite measures have been shown to compare well (22), and were significantly correlated in this study (r=0.874; p<0.001) to the Matsuda Index. Early phase insulin response to the OGTT was estimated using the insulinogenic index (IGI) (23). Participants without data at 30 minutes or whose insulin response was <0 were excluded from the analysis. Insulin secretion was calculated using the C-peptide index, the ratio of the increment in C-peptide relative to glucose in the first 30 minutes of the OGTT (23). C-peptide is produced in equimolar quantities to endogenous insulin, and unlike insulin, there is negligible hepatic extraction of C-peptide, and hence the C-peptide index and the C-peptide to insulin ratio may serve as proxy measures of insulin secretion and clearance, respectively (24,25). Basal and postprandial insulin clearance were calculated as the ratio of fasting C–peptide to insulin, and the incremental area under the curve (iAUC) of C-peptide to iAUC insulin, calculated using the trapezoidal method, respectively. The oral disposition index (oDI), which reflects insulin secretion adjusted for the level of insulin sensitivity (26–28), was calculated as the product of the C-peptide index and Matsuda index (23) which demonstrated a hyperbolic relationship and was used as the measure of beta-cell function. These calculations were only performed in participants without known T2D and/or not taking medications for T2D, and who underwent an OGTT.

### Statistical analysis

Data were analysed using Stata 15.1/IC (StataCorp, College Station, TX, USA). Variables are summarised as percentages for categorical data, mean ± standard deviation (SD) for normally distributed continuous data, and median (25^th^-75^th^ percentile) if not normally distributed. Normality was assessed using the Shapiro–Wilk test and Q-Q probability plots. Sex differences were determined using Students t-test for normally distributed continuous data, Mann-Whitney U and Kruskal–Wallis tests for skewed continuous data, and Chi-squared test for categorical data. Sex differences in glucose and insulin measures are presented before and after adjusting for FMI using one–way analysis of covariance (ANCOVA). Z-scores were derived for the total and regional adiposity measures for the combined sample, as well as sex-stratified using Fisher’s Yates transformation (29). By using Z-scores we were able to compare the risk magnitude per 1 SD change in total and regional adiposity measurements. Multinomial logistic regression was used to explore the relationship between total and regional adiposity measures, and IGM and T2D, using NGT as the reference, and including age, sex, smoking, alcohol intake, education, and FMI (for regional measures), as covariates. All participants with known (n=65) and newly diagnosed (n=42) diabetes were included in the multinomial analyses. We explored sex*adiposity z-score interactions and only found a significant interaction for FMI. Accordingly, the data (excluding FMI) were analysed in the combined sample and the relative risk ratio (RRR) and 95% confidence intervals for IGM and T2D are presented. For the continuous measures of insulin sensitivity (Matsuda index), clearance (fasting C–peptide/insulin ratio) and beta-cell function (oDI), robust regressions were used to explore associations with adiposity z-scores, including age, smoking, alcohol intake, education and FMI (for regional adiposity measures) as covariates. As we were exploring risk factors for T2DM, participants with known diabetes and/or taking medication for diabetes and those without OGTT data were excluded from the robust regression analyses. Due to significant sex interactions in most models, the analyses were completed separately for men and women using sex-specific total and regional adiposity z-scores. A p– value of <0.05 was considered significant.

## Results

### Socio-demographic and body composition characteristics

A total of 804 participants (48.3% men) with a mean age of 54.6±6.0 years were included (Table 1). Men were younger than women and significantly more men were married than women. Current employment status was not different between the sexes, however more men than women (18.1 vs. 12.5%) had completed tertiary education. More men currently smoked (46.1% vs. 7.2%) and frequently consumed alcohol (30.4% vs. 4.6%) than women.

**Table 1:**
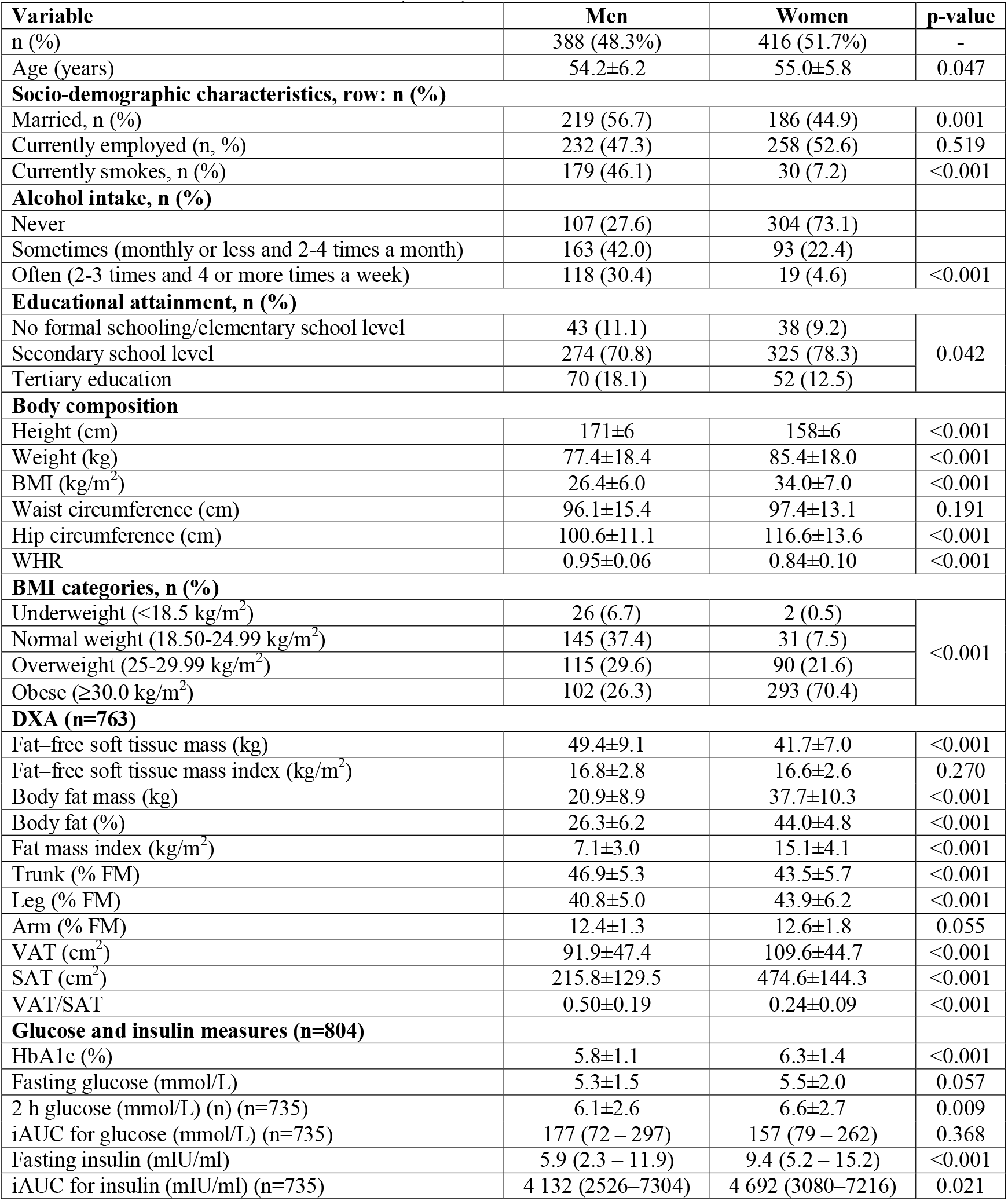
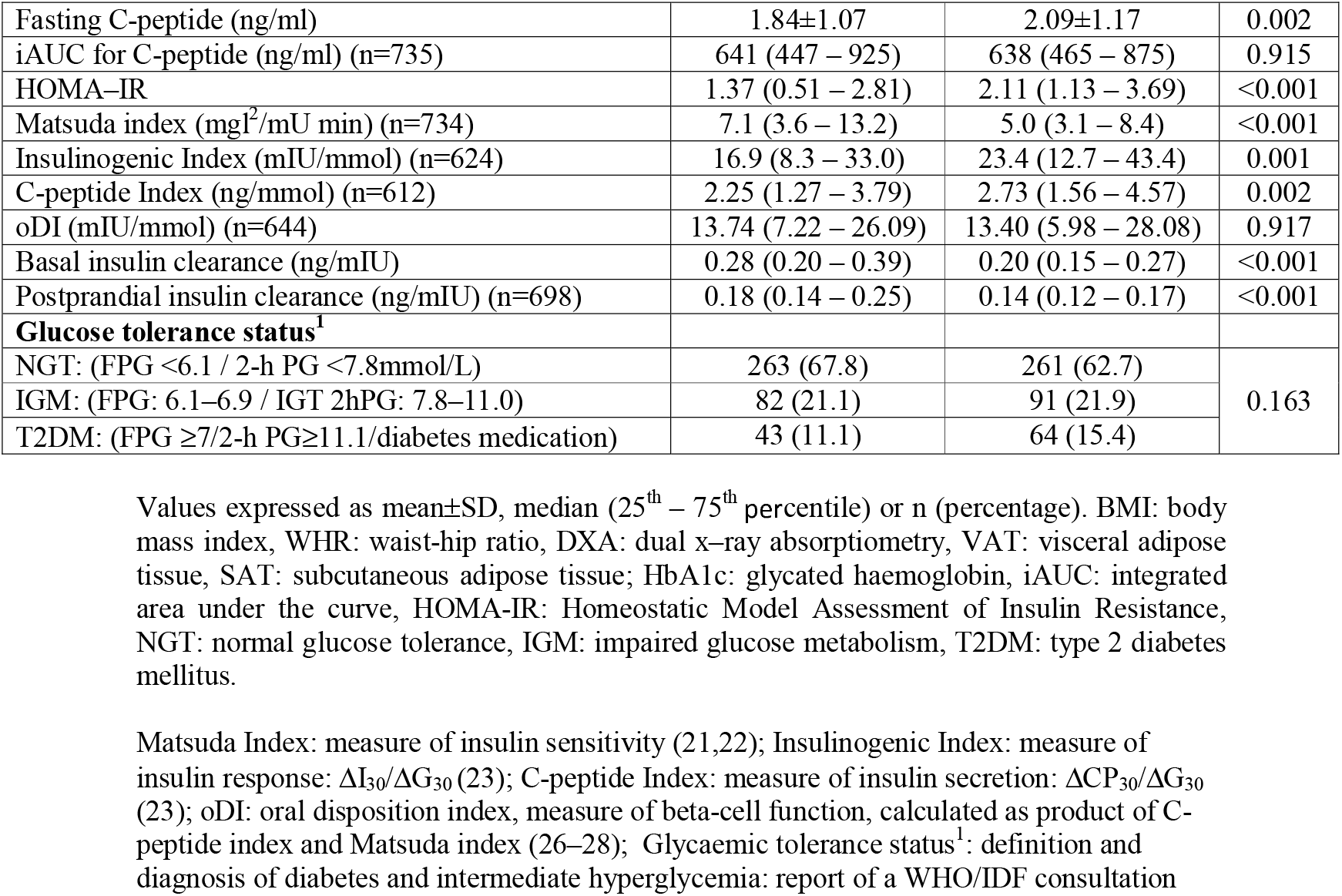
Socio-demographic, body composition, glucose and insulin measures in black South African men and women (n=804)

| Variable | Men | Women | p-value |
| --- | --- | --- | --- |
| n (%) | 388 (48.3%) | 416 (51.7%) | - |
| Age (years) | 54.2±6.2 | 55.0±5.8 | 0.047 |
| Socio-demographic characteristics, row: n (%) |  |  |  |
| Married, n (%) | 219 (56.7) | 186 (44.9) | 0.001 |
| Currently employed (n, %) | 232 (47.3) | 258 (52.6) | 0.519 |
| Currently smokes, n (%) | 179 (46.1) | 30 (7.2) | <0.001 |
| Alcohol intake, n (%) |  |  |  |
| Never | 107 (27.6) | 304 (73.1) |  |
| Sometimes (monthly or less and 2-4 times a month) | 163 (42.0) | 93 (22.4) |  |
| Often (2-3 times and 4 or more times a week) | 118 (30.4) | 19 (4.6) | <0.001 |
| Educational attainment, n (%) |  |  |  |
| No formal schooling/elementary school level | 43 (11.1) | 38 (9.2) | 0.042 |
| Secondary school level | 274 (70.8) | 325 (78.3) |  |
| Tertiary education | 70 (18.1) | 52 (12.5) |  |
| Body composition |  |  |  |
| Height (cm) | 171±6 | 158±6 | <0.001 |
| Weight (kg) | 77.4±18.4 | 85.4±18.0 | <0.001 |
| BMI (kg/m<sup>2</sup>) | 26.4±6.0 | 34.0±7.0 | <0.001 |
| Waist circumference (cm) | 96.1±15.4 | 97.4±13.1 | 0.191 |
| Hip circumference (cm) | 100.6±11.1 | 116.6±13.6 | <0.001 |
| WHR | 0.95±0.06 | 0.84±0.10 | <0.001 |
| BMI categories, n (%) |  |  |  |
| Underweight (<18.5 kg/m<sup>2</sup>) | 26 (6.7) | 2 (0.5) | <0.001 |
| Normal weight (18.50-24.99 kg/m<sup>2</sup>) | 145 (37.4) | 31 (7.5) |  |
| Overweight (25-29.99 kg/m<sup>2</sup>) | 115 (29.6) | 90 (21.6) |  |
| Obese (≥30.0 kg/m<sup>2</sup>) | 102 (26.3) | 293 (70.4) |  |
| DXA (n=763) |  |  |  |
| Fat-free soft tissue mass (kg) | 49.4±9.1 | 41.7±7.0 | <0.001 |
| Fat-free soft tissue mass index (kg/m<sup>2</sup>) | 16.8±2.8 | 16.6±2.6 | 0.270 |
| Body fat mass (kg) | 20.9±8.9 | 37.7±10.3 | <0.001 |
| Body fat (%) | 26.3±6.2 | 44.0±4.8 | <0.001 |
| Fat mass index (kg/m<sup>2</sup>) | 7.1±3.0 | 15.1±4.1 | <0.001 |
| Trunk (% FM) | 46.9±5.3 | 43.5±5.7 | <0.001 |
| Leg (% FM) | 40.8±5.0 | 43.9±6.2 | <0.001 |
| Arm (% FM) | 12.4±1.3 | 12.6±1.8 | 0.055 |
| VAT (cm<sup>2</sup>) | 91.9±47.4 | 109.6±44.7 | <0.001 |
| SAT (cm<sup>2</sup>) | 215.8±129.5 | 474.6±144.3 | <0.001 |
| VAT/SAT | 0.50±0.19 | 0.24±0.09 | <0.001 |
| Glucose and insulin measures (n=804) |  |  |  |
| HbA1c (%) | 5.8±1.1 | 6.3±1.4 | <0.001 |
| Fasting glucose (mmol/L) | 5.3±1.5 | 5.5±2.0 | 0.057 |
| 2 h glucose (mmol/L) (n) (n=735) | 6.1±2.6 | 6.6±2.7 | 0.009 |
| iAUC for glucose (mmol/L) (n=735) | 177 (72 – 297) | 157 (79 – 262) | 0.368 |
| Fasting insulin (mIU/ml) | 5.9 (2.3 – 11.9) | 9.4 (5.2 – 15.2) | <0.001 |
| iAUC for insulin (mIU/ml) (n=735) | 4 132 (2526–7304) | 4 692 (3080–7216) | 0.021 |
| Fasting C-peptide (ng/ml) | 1.84±1.07 | 2.09±1.17 | 0.002 |
| iAUC for C-peptide (ng/ml) (n=735) | 641 (447 – 925) | 638 (465 – 875) | 0.915 |
| HOMA-IR | 1.37 (0.51 – 2.81) | 2.11 (1.13 – 3.69) | <0.001 |
| Matsuda index ($\text{mg l}^2/\text{mU min}$) (n=734) | 7.1 (3.6 – 13.2) | 5.0 (3.1 – 8.4) | <0.001 |
| Insulinogenic Index (mIU/mmol) (n=624) | 16.9 (8.3 – 33.0) | 23.4 (12.7 – 43.4) | 0.001 |
| C-peptide Index (ng/mmol) (n=612) | 2.25 (1.27 – 3.79) | 2.73 (1.56 – 4.57) | 0.002 |
| oDI (mIU/mmol) (n=644) | 13.74 (7.22 – 26.09) | 13.40 (5.98 – 28.08) | 0.917 |
| Basal insulin clearance (ng/mIU) | 0.28 (0.20 – 0.39) | 0.20 (0.15 – 0.27) | <0.001 |
| Postprandial insulin clearance (ng/mIU) (n=698) | 0.18 (0.14 – 0.25) | 0.14 (0.12 – 0.17) | <0.001 |
| **Glucose tolerance status<sup>1</sup>** |  |  |  |
| NGT: (FPG <6.1 / 2-h PG <7.8mmol/L) | 263 (67.8) | 261 (62.7) | 0.163 |
| IGM: (FPG: 6.1–6.9 / IGT 2hPG: 7.8–11.0) | 82 (21.1) | 91 (21.9) |  |
| T2DM: (FPG $\geq 7/2$-h PG $\geq 11.1$/diabetes medication) | 43 (11.1) | 64 (15.4) | |

Values expressed as mean±SD, median (25<sup>th</sup> – 75<sup>th</sup> percentile) or n (percentage). BMI: body mass index, WHR: waist-hip ratio, DXA: dual x-ray absorptiometry, VAT: visceral adipose tissue, SAT: subcutaneous adipose tissue; HbA1c: glycated haemoglobin, iAUC: integrated area under the curve, HOMA-IR: Homeostatic Model Assessment of Insulin Resistance, NGT: normal glucose tolerance, IGM: impaired glucose metabolism, T2DM: type 2 diabetes mellitus.
Matsuda Index: measure of insulin sensitivity (21,22); Insulinogenic Index: measure of insulin response: $\Delta I_{30}/\Delta G_{30}$ (23); C-peptide Index: measure of insulin secretion: $\Delta CP_{30}/\Delta G_{30}$ (23); oDI: oral disposition index, measure of beta-cell function, calculated as product of C-peptide index and Matsuda index (26–28); Glycaemic tolerance status<sup>1</sup>: definition and diagnosis of diabetes and intermediate hyperglycemia: report of a WHO/IDF consultation

Mean BMI was higher in women than men (p<0.001), and accordingly a larger proportion of the women presented with obesity (70.2% vs. 26.6%) (Table 1). While waist circumference was similar, men had higher WHR due to the higher hip circumference of the women. While FFSTM was higher in men, FM (kg and %) and FMI were higher in women. When expressed relative to FM, women had significantly greater leg FM, while men had more central FM (trunk), but arm FM did not differ. Within the central depot, men had less VAT and SAT (both p<0.001), but a higher VAT/SAT ratio.

### Differences in glucose and insulin measures between men and women

Although fasting glucose and iAUC for glucose were not different between the sexes, HbA1C and 2 h glucose were higher in women than men (Table 1). Fasting insulin and C-peptide, and iAUC for insulin, were also higher in women than men. Accordingly, HOMA-IR was higher and insulin sensitivity (Matsuda index) was lower in women compared to men, accompanied by a higher insulin response (IGI) characterised by higher insulin secretion (C-peptide index) and lower insulin clearance (basal and postprandial). However, the oDI, a measure of beta-cell function, did not differ by sex.

When adjusting for differences in FMI (Table 2), there were no longer sex differences in HbA1C, 2-hour glucose, insulin response, or basal and postprandial insulin clearance, while insulin secretion remained higher in women. In contrast, fasting insulin and C-peptide, as well as HOMA-IR were higher, and insulin sensitivity and beta-cell function were lower in men compared to women.

**Table 2:**
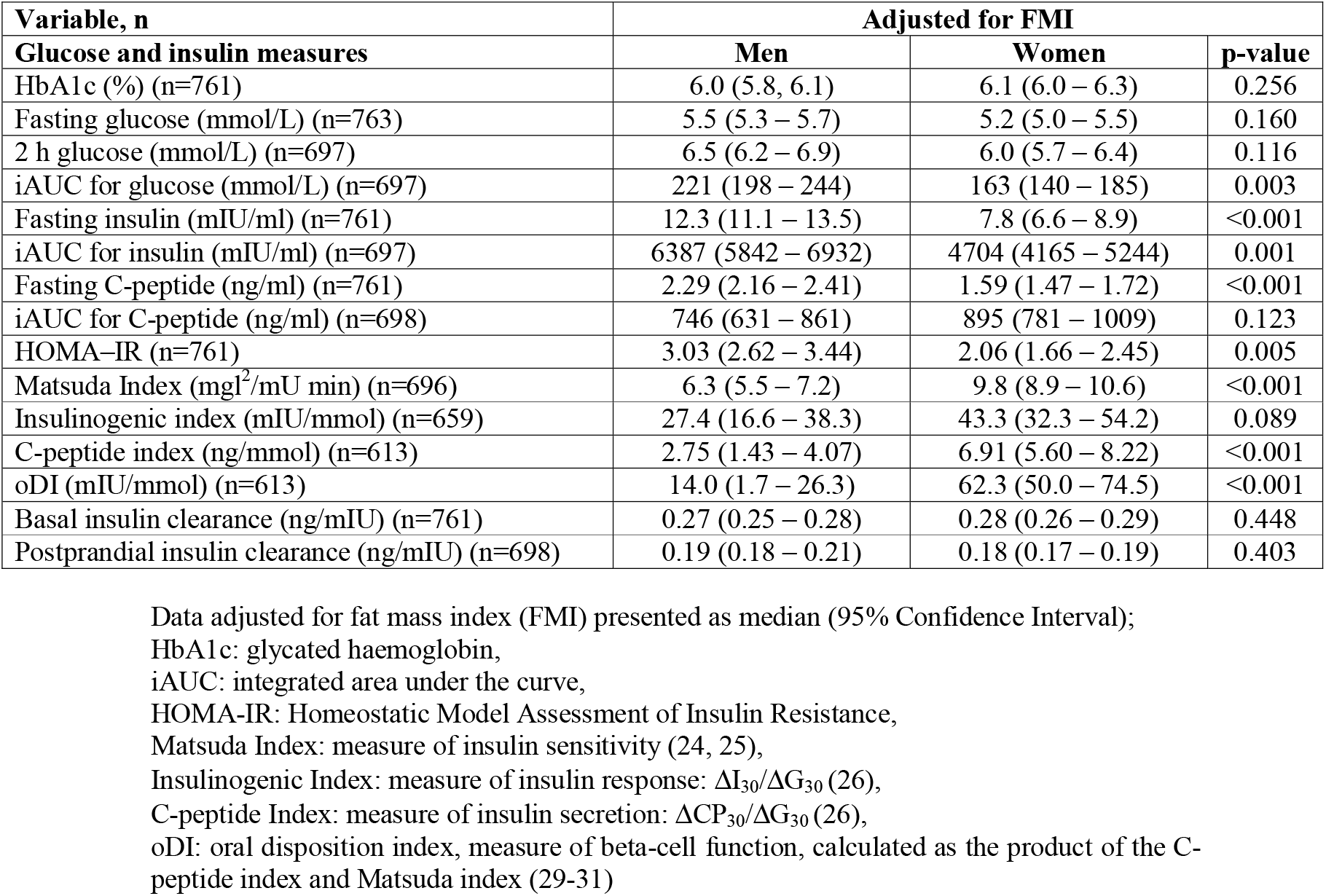
Glucose and insulin measures in black SA men and women adjusted for FMI

The prevalences of NGT, IGM and T2DM were not significantly different between men and women.

### Associations between total and regional adiposity and risk for IGM and type 2 diabetes

There was a significant sex*FMI z-score interaction (p<0.001), such that the RRR for IGM and T2D were greater for men than women (Figure 1A). Associations between regional adiposity z-scores and risk for IGM and T2D did not differ by sex, and the RRR for the combined sample are presented in Table 3. Trunk fat and VAT z-scores were associated with a higher risk for both IGM and T2D, with every 1 SD increase in trunk fat and VAT being associated with a 4.8 fold and 2.6 fold increased risk for T2D, respectively. In contrast, higher leg fat z-score was associated with a 58% and 79% lower risk for IGM and T2D, respectively, while a 1 SD higher arm fat z-score was associated with a 2.2-fold greater risk for T2D only. SAT z-score was not associated with IGM or T2D.

**Table 3:** Associations between regional adiposity z-scores and risk for IGM and type 2 diabetes in men and women combined

| **Men and women** | **RRR** | **95% CI** | **p-value** | **Model R<sup>2</sup>** |
| --- | --- | --- | --- | --- |
| **Trunk z-score (n=761)** |  |  |  |  |
| IGM | 2.35 | 1.43 – 3.87 | 0.001 | <0.001 |
| Type 2 diabetes | 4.76 | 2.68 – 8.45 | <0.001 |  |
| **Leg z-score (n=759)** |  |  |  |  |
| IGM | 0.42 | 0.24 – 0.71 | 0.001 | <0.001 |
| Type 2 diabetes | 0.21 | 0.11 – 0.39 | <0.001 |  |
| **Arm z-score (n=759)** |  |  |  |  |
| IGM | 1.35 | 0.83 – 2.21 | 0.224 | <0.001 |
| Type 2 diabetes | 2.19 | 1.25 – 3.81 | 0.006 |  |
| **VAT z-score (n=753)** |  |  |  |  |
| IGM | 1.76 | 1.35 – 2.28 | <0.001 | <0.001 |
| Type 2 diabetes | 2.58 | 1.92 – 3.48 | <0.001 |  |
| **SAT z-score (n=753)** |  |  |  |  |
| IGM | 1.35 | 0.74 – 2.48 | 0.331 | <0.001 |
| Type 2 diabetes | 1.37 | 0.68 – 2.75 | 0.376 |  |

Results of multinomial logistic regression presented as relative risk ratios (RRR) and 95% confidence interval and represent risk of outcome with 1 SD increase in regional adiposity. Model used normal glucose tolerance (NGT) as the reference group compared to impaired glucose metabolism (IGM) and type 2 diabetes, adjusted for: age, smoking, alcohol intake, education attainment, FMI and sex

### Sex-specific associations between total and regional adiposity z-scores and insulin measures

There were significant sex*FMI z-score interactions for insulin sensitivity, clearance and beta-cell function, with associations consistently being stronger in men than women (Figure 1B–1D). There were also significant sex*regional adiposity interactions for most measures of insulin sensitivity and response and therefore the results are presented separately for men and women (Table 4). Lower insulin sensitivity was associated with higher central fat mass (trunk fat and VAT), and lower leg fat in both men and women, but the associations with central fat mass were stronger in men than women (p<0.001 for all interactions). In contrast, arm fat mass was associated with lower insulin sensitivity in women only (p<0.001 for interaction). Beta-cell function (oDI) was negatively associated with VAT in both men and women. In contrast beta-cell function was positively associated with peripheral fat mass in women only (p=0.040 for interaction). Basal insulin clearance was negatively associated with trunk fat mass in both men and women, with a stronger association in men (p=0.017 for interaction). In contrast, basal insulin clearance was negatively associated with VAT and arm fat, and positively associated with leg fat in women only, but the strength of the association did not differ significantly between sexes. The associations for postprandial insulin clearance were similar to those for basal insulin clearance (data not shown). As the women were at different phases of the menopausal transition with 17.6% being premenopausal, 14.7% perimenopausal and 67.7% being postmenopausal, we wanted to ascertain whether the associations presented above differed by menopausal phase. The associations between total and regional adiposity and insulin sensitivity, secretion and beta-cell function did not differ between menopausal groups. In contrast, the associations between FMI, trunk, leg and arm z-scores and basal insulin clearance differed by menopausal phase, being stronger in the pre-than peri- and postmenopausal women (data not shown).

**Table 4:** Associations between regional adiposity z-scores and insulin sensitivity, basal insulin clearance and Beta-cell function

| | $\beta$ | 95% CI | p-value | Model R<sup>2</sup> | Model p-value | $\beta$ | 95% CI | p-value | Model R<sup>2</sup> | Model p-value |
| --- | --- | --- | --- | --- | --- | --- | --- | --- | --- | --- |
|  | **Men** |  |  |  |  | **Women** |  |  |  |  |
|  | **Insulin Sensitivity (n=344)** |  |  |  |  | **Insulin Sensitivity (n=349)** |  |  |  |  |
| FMI z-score<sup>#</sup> | -3.442 | -4.011 to -2.873 | <0.001 | 0.284 | <0.001 | -1.225 | -1.631 to -0.819 | <0.001 | 0.069 | <0.001 |
| Trunk z-score<sup>#</sup> | -2.680 | -4.829 to -0.532 | 0.015 | 0.295 | <0.001 | -1.748 | -2.545 to -0.950 | <0.001 | 0.097 | <0.001 |
| Leg z-score | 1.695 | 0.233 to 3.157 | 0.023 | 0.293 | <0.001 | 1.218 | 0.454 to 1.982 | 0.002 | 0.086 | <0.001 |
| Arm z-score<sup>#</sup> | 0.027 | -1.667 to 1.722 | 0.975 | 0.284 | <0.001 | -0.893 | -1.716 to -0.070 | 0.033 | 0.080 | <0.001 |
| VAT z-score<sup>#</sup> | -1.679 | -2.540 to -0.818 | <0.001 | 0.309 | <0.001 | -1.174 | -1.657 to -0.691 | <0.001 | 0.106 | <0.001 |
| SAT z-score<sup>#</sup> | -1.043 | -2.961 to 0.876 | 0.286 | 0.286 | <0.001 | -0.693 | -1.582 to 0.195 | 0.126 | 0.075 | <0.001 |
|  | **Beta-cell function (n=304)** |  |  |  |  | **Beta-cell function (n=306)** |  |  |  |  |
| FMI z-score<sup>#</sup> | -3.9594 | -5.3600 - -2.5589 | <0.001 | 0.065 | <0.001 | -2.6420 | -4.0684 to -1.2156 | <0.001 | 0.027 | 0.001 |
| Trunk z-score | -4.0597 | -9.2112 to 1.0917 | 0.122 | 0.068 | <0.001 | -4.3905 | -7.2286 to -1.5524 | 0.003 | 0.040 | <0.001 |
| Leg z-score<sup>#</sup> | 3.182 | -0.3294 to 6.6940 | 0.076 | 0.071 | <0.001 | 4.3120 | 1.7781 to 6.8460 | 0.001 | 0.044 | <0.001 |
| Arm z-score<sup>#</sup> | 0.1387 | -3.8845 to 4.1618 | 0.946 | 0.065 | <0.001 | -0.5693 | -3.3668 to 2.2282 | 0.689 | 0.028 | 0.001 |
| VAT z-score | -2.6180 | -4.7016 to 0.5344 | 0.014 | 0.074 | <0.001 | -3.8567 | -5.5820 to -2.1314 | <0.001 | 0.051 | <0.001 |
| SAT z-score | 1.6166 | -2.9625 to 6.1957 | 0.488 | 0.066 | <0.001 | -1.8141 | -4.7217 to 1.0936 | 0.220 | 0.029 | 0.001 |
|  | **Basal Insulin Clearance (n=346)** |  |  |  |  | **Basal Insulin Clearance (n=351)** |  |  |  |  |
| FMI z-score<sup>#</sup> | -0.047 | -0.062 to -0.033 | <0.001 | 0.121 | <0.001 | -0.027 | -0.035 to -0.018 | <0.001 | 0.092 | <0.001 |
| Trunk z-score<sup>#</sup> | -0.055 | -0.109 to -0.001 | 0.047 | 0.129 | <0.001 | -0.026 | -0.044 to -0.009 | 0.002 | 0.108 | <0.001 |
| Leg z-score | 0.006 | -0.031 to 0.044 | 0.738 | 0.121 | <0.001 | 0.017 | 0.001 to 0.033 | 0.036 | 0.099 | <0.001 |
| Arm z-score<sup>#</sup> | -0.015 | -0.058 to 0.027 | 0.477 | 0.122 | <0.001 | -0.022 | -0.038 to -0.005 | 0.011 | 0.104 | <0.001 |
| VAT z-score | 0.001 | -0.022 to 0.023 | 0.945 | 0.121 | <0.001 | -0.018 | -0.028 to -0.008 | 0.001 | 0.111 | <0.001 |
| SAT z-score<sup>#</sup> | -0.033 | -0.082 to 0.015 | 0.175 | 0.125 | <0.001 | -0.005 | -0.024 to 0.013 | 0.555 | 0.093 | <0.001 |

Beta coefficients for robust regression models for men and women, adjusted for age, smoking, alcohol intake, education attainment and FMI (except for FMI z-score), VAT: visceral adipose tissue, SAT: subcutaneous adipose tissue, Insulin sensitivity estimated from the Matsuda Index (24, 25) and beta-cell function was estimated using the oral Disposition index calculated as the product of C-peptide and Matsuda Index (26), and basal insulin clearance calculated as the ratio of fasting C-peptide to fasting insulin
<sup>#</sup>p<0.05 for sex\*z-score body fat interaction term

## Discussion

The main and novel findings of this study were that in a sample of black men and women with a mean age of 54.6 years, after adjustments for differences in body fat, insulin sensitivity, secretion and beta-cell function were lower in black SA men compared to women, while insulin clearance did not differ by sex. In line with this, the strength of the association between total adiposity and T2D risk was greater in men compared to women. Although black SA women have a higher prevalence of obesity (70.2 vs. 26.6%) and greater whole-body fatness (43.6 vs. 26.3%) than men, they present with a more ‘favourable’ body fat distribution, characterised by less central fat mass and greater peripheral fat mass. This phenotype has been associated with lower diabetes risk. This together with the greater impact of body fatness on diabetes risk could explain the similar prevalence of diabetes in men and women (11.1 vs. 15.4%) despite the lower adiposity in men in this study.

These findings also suggest that with increasing adiposity, black SA men will be at greater risk for T2D than their female counterparts. We found that the association between total adiposity and risk for T2D was higher in men than women (Figure 1A). Further we showed that with increasing FMI the decline in insulin sensitivity was greater in men compared to women, similar to earlier studies from SA (8,30), which was also associated with a more pronounced decrease in beta-cell function in men compared to women. The higher risk in men compared to women was independent of smoking and alcohol intake, lifestyle risk factors were also higher in men compared to women.

The finding of similar T2D prevalence (1) despite marked differences in the prevalence of obesity (3) between sexes are consistent and representative of South Africa and the SSA region. In order to understand the sexual dimorphism in this relationship, it is obviously essential to account for sex differences in body fatness as well as disentangle the sex-specific associations between regional adiposity and T2D risk. After adjusting for differences in body fatness, men had lower insulin sensitivity, insulin secretion and beta-cell function compared to women, placing the men at higher risk for T2D. Indeed, a lower beta-cell function, estimated using the oDI, has been shown to predict the development of T2D over a 10 year period in a Japanese American cohort (26).

Black African women have been shown to present with hyperinsulinaemia compared to their European counterparts, often beyond that required to maintain normoglycaemia (8,31). Hyperinsulinemia in black African women has previously been attributed to alterations in both insulin secretion and clearance, depending on age, and/or level of glycemia (10,32). Studies in African American women have shown that decreased hepatic insulin clearance is the main contributor to hyperinsulinemia (33). In contrast, we show that the higher IGI in women compared to men, was associated with higher insulin secretion without differences in insulin clearance. Due to limited longitudinal studies, it is not known whether the higher IGI in women is protective or may actually cause insulin resistance (9).

It is well recognised globally and in South Africa that men have greater central body fat and less lower body peripheral fat compared to women (8,30). Similarly, we showed that men had greater trunk fat mass, a higher VAT/SAT ratio, and less leg and similar arm fat mass than women. This adiposity phenotype is associated with greater diabetes risk as previously reported by our group (8,30,32,34). Indeed, we showed that a 1SD increase in trunk z-score was associated with a more than two-fold greater risk for IGM and nearly five times greater risk for T2D, and was also associated with lower insulin sensitivity and lower basal insulin clearance. In contrast, peripheral fat is typically associated with reduced risk for diabetes (30,34) as it acts as a metabolic sink to sequester excess free fatty acids that may otherwise be directed at ectopic sites such as the liver and pancreas (35). We showed that a 1 SD increase in leg z-score was associated with a 58% lower risk for IGM and a 79% lower risk for T2D, as well as higher insulin sensitivity in both sexes. Notably, the strength of the inverse association between central fat distribution and insulin sensitivity was greater in men compared to women. Several studies in different populations have shown VAT to be more strongly associated with insulin resistance, and therefore a greater risk for T2DM, in men than women (30,36–38). A further novel finding of the study was that the positive relationship between beta-cell function and leg FM was weaker in men compared to women, suggesting lower ‘protective’ effect of leg FM on beta-cell function in men compared to women. Accordingly, despite a lower prevalence of overweight and obesity in men compared to women in our study, this ‘unfavourable’ regional fat distribution and the sex-specific relationships with insulin sensitivity and beta-cell function places them at greater risk for future T2D.

This is the first study, to our knowledge, in black SA men and women with detailed measures of insulin sensitivity, secretion and clearance, and beta-cell function, based on estimates from an OGTT. We were also able to use DXA, which provides an accurate assessment of body composition and regional adiposity. A limitation is the cross–sectional nature of the study which does not allow us to infer causality. Although the sex differences in obesity and total adiposity may be seen as a limitation, it reflects the status of obesity within South Africa and the sub-Saharan African region (3), and adjustments for total body fatness and the calculation of z-scores were used in the analyses to determine whether these sex differences in adiposity were influencing the insulin- and glucose-related variables. There were no effects of the menopausal transition *per se* on the association between adiposity and insulin sensitivity, secretion and beta-cell function and therefore the sex differences reported cannot be explained by menopausal status. However, the premenopausal women were not tested at a specific time during their menstrual cycle, which is noted as a limitation of the study. Furthermore, the conclusions for this study are valid only for HIV negative individuals. In summary, for the same level of body fatness, black South African men are less insulin sensitive and had lower insulin secretion and beta-cell function than women, with the strength of the association between adiposity and T2D risk being greater in men compared to women. This suggests that with increasing adiposity, particularly an increase in central adiposity, black SA men face an increased risk for T2D in comparison with their female counterparts. Longitudinal studies are required to confirm the results of this study.

## Data Availability

All data produced in the present study are available upon reasonable request to the authors

## Funding

The study was jointly funded by the South African Medical Research Council (MRC) from South African National Department of Health, MRC UK (via the Newton Fund) and GSK Africa Non-Communicable Disease Open Lab (via a supporting Grant project Number: ES/N013891/1) and South African National Research Foundation (Grant no: UID:98561).

## Conflict of Interest

No potential conflicts of interest relevant to this article were reported

## Authors’ contributions

CNK, JHG and LKM designed the study and CNK analysed the data, drafted and revised the manuscript under the supervision of JHG and LKM. All authors reviewed/edited, read and approved all the drafts and the final version of the manuscript. The authors retained the control of the final content of the publication.

## Acknowledgements

We are grateful to the participants as well as the following DPHRU staff for their input during data collection and entry: Melikhanya Soboyisi, Tshifhiwa Ratshikombo, Vukosi Mkansi, Sphume Thango, Mosadiapula Nakedi, Thabile Sibiya, Bonisiwe Mlambo, Caroline Makura, Dr Mamosilo Lichaba, Karabo Pearl Nkhahle and team, and Prof Michèle Ramsay for AWI-Gen study development and oversight.

## Notes

### Competing Interest Statement

The authors have declared no competing interest.

### Author Declarations

The study was approved by the Human Research Ethics Committee (HREC) of the University of the Witwatersrand (M160604 and M160975), Johannesburg, South Africa.

